# Employing a Systematic Approach to Biobanking and Analyzing Clinical and Genetic Data for Advancing COVID-19 Research

**DOI:** 10.1101/2020.07.24.20161307

**Authors:** Sergio Daga, Chiara Fallerini, Margherita Baldassarri, Francesca Fava, Floriana Valentino, Gabriella Doddato, Elisa Benetti, Simone Furini, Annarita Giliberti, Rossella Tita, Sara Amitrano, Mirella Bruttini, Ilaria Meloni, Anna Maria Pinto, Francesco Raimondi, Alessandra Stella, Filippo Biscarini, Nicola Picchiotti, Marco Gori, Pietro Pinoli, Stefano Ceri, Maurizio Sanarico, Francis P. Crawley, GEN-COVID Multicenter Study, Alessandra Renieri, Francesca Mari, Elisa Frullanti

**Affiliations:** Medical Genetics, University of Siena, Italy; Genetica Medica, Azienda Ospedaliero-Universitaria Senese, Italy; Department of Medical Biotechnologies, University of Siena, Italy; Scuola Normale Superiore, Pisa, Italy; CNR-Consiglio Nazionale delle Ricerche, Istituto di Biologia e Biotecnologia Agraria (IBBA), Milano, Italy; University of Siena, DIISM-SAILAB, Siena, Italy; Department of Mathematics, University of Pavia, Pavia, Italy; Université Côte d’Azur, Inria, CNRS, I3S, Maasai; Department of Electronics, Information and Bioengineering (DEIB), Politecnico di Milano, Milano, Italy; Independent Data Scientist, Milan, Italy; Good Clinical Practice Alliance-Europe (GCPA) and Strategic Initiative for Developing Capacity in Ethical Review-Europe (SIDCER), Brussels, Belgium

**Keywords:** COVID-19, Biobank, Registry, WES, GWAS

## Abstract

Within the GEN-COVID Multicenter Study, biospecimens from more than 1,000 SARS-CoV-2-positive individuals have thus far been collected in the GEN-COVID Biobank (GCB). Sample types include whole blood, plasma, serum, leukocytes, and DNA. The GCB links samples to detailed clinical data available in the GEN-COVID Patient Registry (GCPR). It includes hospitalized patients (74.25%), broken down into intubated, treated by CPAP-biPAP, treated with O2 supplementation, and without respiratory support (9.5%, 18.4%, 31.55% and 14.8, respectively); and non-hospitalized subjects (25.75%), either pauci- or asymptomatic. More than 150 clinical patient-level data fields have been collected and binarized for further statistics according to the organs/systems primarily affected by COVID-19: heart, liver, pancreas, kidney, chemosensors, innate or adaptive immunity, and clotting system. Hierarchical Clustering analysis identified five main clinical categories: i) severe multisystemic failure with either thromboembolic or pancreatic variant; ii) cytokine storm type, either severe with liver involvement or moderate; iii) moderate heart type, either with or without liver damage; iv) moderate multisystemic involvement, either with or without liver damage; v) mild, either with or without hyposmia. GCB and GCPR are further linked to the GEN-COVID Genetic Data Repository (GCGDR), which includes data from Whole Exome Sequencing and high-density SNP genotyping. The data are available for sharing through the Network for Italian Genomes, found within the COVID-19 dedicated section. The study objective is to systematize this comprehensive data collection and begin identifying multi-organ involvement in COVID-19, defining genetic parameters for infection susceptibility within the population and mapping genetically COVID-19 severity and clinical complexity among patients.

## INTRODUCTION

The GEN-COVID Multicenter Study was designed to collect and systematize biological samples and clinical data across multiple hospitals and healthcare facilities in Italy with the purpose of deriving patient-level phenotypic and genotypic data and the specific intention to make samples and data available to COVID-19 researchers globally. To reach these aims, the project collected and organized high-quality samples and data whose integrity was assured and could be readily accessed and processed for COVID-19 research using existing interoperability standards and tools. To this end, a GEN-COVID Biobank (GCB) and a GEN-COVID Patient Registry (GCPR) were established utilizing already existing biobanking and patient registry infrastructure. The collection of samples and data are now utilized in the GEN-COVID Multicenter Study for generating Genotyping (GWAS) and Whole Exome Sequencing (WES) results. This study also works collaboratively with other genomic studies on COVID-19. The data resulting from these studies is then stored and made available through the GEN-COVID Genetic Data Repository (GCGDR). All samples and data have also been systematized in accordance with the FAIR (Findability, Accessibility, Interoperability, and Reuse) Data Principles [1] to promote their international availability and use for COVID-19 research.

The outbreak of the coronavirus disease 2019 (COVID-19), the Severe Acute Respiratory Syndrome caused by coronavirus SARS-CoV-2, that first appeared in December 2019 in Wuhan, Huanan, Hubei Province of China, has resulted in millions of cases worldwide within a few short months, and rapidly evolved into a real pandemic [2]. The COVID-19 pandemic represents an enormous challenge to the world’s healthcare systems. Among the European countries, Italy was the first to experience the epidemic wave of SARS-CoV-2 infection, accompanied by a severe clinical picture and a mortality rate reaching 14%. In Italy, as of July 16th, 2020, there were 243,506 confirmed COVID-19 cases and 34,997 related deaths reported [3].

The disease is characterized by a highly heterogeneous phenotypic response to SARS-CoV-2 infection, with the large majority of infected individuals having only mild or even no symptoms. However, the severe cases can rapidly evolve towards a critical respiratory distress syndrome and multiple organ failure. The symptoms of COVID-19 range from fever, cough, sore throat, congestion, and fatigue to shortness of breath, hemoptysis, pneumonia followed by respiratory disorders and septic shock [4].

The overburdened healthcare infrastructure and the working conditions within healthcare centers are tremendously challenging. Direct patient care is given the highest priority. Focus is concentrated on monitoring infection evolution in terms of the number of new cases and the number of deaths. Disease severity is also an important parameter that is being continually evaluated, with a current focus on patients experiencing serious pulmonary disease and other life-threatening conditions. Although patient care is the first priority, in the public health emergency situation brought on by the COVID-19 pandemic, it is also of the utmost importance to collect, process, and share with rapidity and confidence human biological materials, clinical data, and study outcomes. The best suited tool to address this need and accelerate research on COVID-19 is an accessible, high quality biobank with associated clinical data and the necessary tools to guarantee interoperability with other biobanks and databanks.

This paper addresses the main aim of the project: the collection and systematization of human biological materials, clinical data stored in a patient registry, and derived patient-level genetic data. The paper addresses the methods for sample and data collection, and the systematization of the samples and data for research purposes. As COVID-19 increasingly reveals itself as a multi-systemic disease, the purpose of this data collection is to include the most relevant clinical variables that identify multi-organ involvement as well as identify the genetic determinants of virus-host interaction, so as to holistically disclose the effect of COVID-19 over several physiological subsystems. In the present paper, the samples and the complete datasets are then used within the GEN-COVID Multicenter Study for identifying multi-organ involvement in COVID-19, defining genetic parameters for infection susceptibility within the population, and mapping genetically COVID-19 severity and clinical complexity among patients. Going forward, the main challenge will be to define the genetic parameters for infection susceptibility within specific populations in order to be able to map and identify genetically COVID-19 severity and clinical complexity within and across patient groups.

## METHODS

### Study design

The purpose of the GEN-COVID Multicenter Study is to make the best use of the widest possible sets of patient data and genetic material in order to identify potential links between patient genetic variation and clinical variability, patient presentation and disease severity. By exposing the potential links between genetic variability and disease variability, the study believes it can contribute to improved patient-level diagnostics, prognosis, and personalized treatment of COVID-19. To achieve this overall aim, the following specific objectives are being pursued: i) to perform sequencing (WES) on 2,000 COVID-19 patient samples [performed by the University of Siena (UNISI)]; ii) to perform genotyping (GWAS) on 2000 COVID-19 patients [performed by the Institute for Molecular Medicine of Finland (FIMM)]; iii) to associate the host genetic data obtained on 2,000 COVID-19 patients with severity and prognosis; iv) to share phenotypic data and samples across the GEN-COVID consortium platform as well as in cooperation with research institutions and national platforms through the GEN-COVID Disease Registry and Biobank; v) to share genetic data through the Network of Italian Genome (NIG: http://www.nig.cineca.it/, NIG database: http://nigdb.cineca.it) at CINECA, the largest Italian computing center.

Planned key deliverables of the project are i) to develop a state-of-the-art Patient Registry and Biobank for COVID-19 clinical research with access for academic and industry partners; ii) to understand the genetic and molecular basis of susceptibility to SARS-CoV-2 infection and (susceptibility to a potentially more severe clinical outcome [prognosis] within 12 months); and iii) to understand the genetic profile of patients, contributing to the rapid identification of medicines to be repurposed for personalized therapeutic approaches that demonstrate greater efficacy against the COVID-19 virus. As the initial starting point of this process, the *ACE2* gene has already been extensively investigated in the Italian population [5].

The GEN-COVID Multicenter Study includes a network of 22 Italian hospitals, 13 of which from Northern Italy, 5 from Central Italy, and 4 from Southern Italy. It also includes local healthcare units and departments of preventative medicine (https://sites.google.com/dbm.unisi.it/gen-covid). The network continues to grow as more hospitals and healthcare centers express an interest in contributing samples and data. It started its activity on March 16, 2020, following approval by the Ethical Review Board of the Promoter Center, University of Siena (Protocol n. 16929, approval dated March 16, 2020). Written informed consent was obtained from all individuals who contributed samples and data. Detailed clinical and laboratory characteristics (data), specifically related to COVID-19, were collected for all subjects.

### Study participants and recruitment

In order to ensure a collection that could be, as much as possible, comprehensive and representative of the Italian population, hospitals from across Italy, local healthcare units, and departments of preventative medicine have been involved in collecting samples and associated patient-level data for the GEN-COVID Multicenter Study. The inclusion criteria for the study are PCR-positive SARS-CoV-2 `infection, age ≥ 18 years, and appropriately given informed consent.` In addition to the samples collection, an extensive questionnaire is used to assess disease severity and collect basic demographic information from each patient (**Supplementary Table 1**).

As of July 16th, 2020, we have collected samples and data from 1,033 individuals (1021 without family ties and 12 with family relations). All were positively diagnosed with SARS-CoV-2 and they represent a wide range of disease severity, ranging from hospitalized patients with severe COVID-19 disease to asymptomatic individuals. Infection status was confirmed by SARS-CoV-2 viral RNA polymerase-chain-reaction (PCR) test collected mainly from nasopharyngeal swabs. Recruitment remains ongoing with the goal of including samples and data from 2000 individuals by the end of September 2020. So far recruiting an averages of 200 patients per week.

### Data collection and storage

The GEN-COVID registry was designed in order to guarantee data accuracy and, at the same time, to ensure ease of data entry in order to facilitate compliance and save clinicians time. The highest data integrity and data privacy standards, with reference to the EU General Data Protection Regulation (GDPR) [6], were also built into the training for personnel. Samples and data were collected and systematized in order to meet the FAIR Data Principles requirements.

The socio-demographic information included sex, age, and ethnicity. Information about family history, (pre-existing) chronic conditions, and SARS-CoV-2 related symptoms were collected through a detailed core clinical questionnaire as previously reported [7]. This clinical data was continually updated accordingly as new information appeared regarding COVID-19 (**Supplementary Table 1**). More than 150 clinical items have been collected and synthesized in a binary mode for each involved organ/system: heart, liver, pancreas, kidney, and olfactory/gustatory and lymphoid systems. The collection and organizing methodologies allowed for rapid statistical analysis. Data were handled and stored in accordance with the EU GDPR [6].

Peripheral blood samples in ethylenediamine tetraacetic acid (EDTA)-containing tubes were collected for all subjects. Genomic DNA was centrally isolated from peripheral blood samples using the MagCore®Genomic DNA Whole Blood Kit (Diatech Pharmacogenetics, Jesi, Italy) according to the manufacturer’s protocol at the Promoter Center. For all subjects, aliquots of plasma and serum are also available. Whenever possible, leukocytes were isolated from whole blood by density gradient centrifugation and stored in dimethyl sulfoxide (DMSO) solution and frozen using liquid nitrogen. For the majority of cohort, swab specimens are also available and stored at the reference hospitals.

Genetic data from GWAS and WES were generated for all patients. The generation of such a massive amount of sequencing data required sufficient computing resources able to store and analyse large quantities of data. For this purpose, GEN-COVID took advantage of University of Siena’s participation in the Network for Italian Genomes (NIG, http://www.nig.cineca.it/, NIG database, http://nigdb.cineca.it/), which collects genome sequencing data from the Italian population. NIG has a specific agreement with CINECA, the largest computing centre in Italy and one of the largest in Europe, for the use of the CINECA facility for the storage and analysis of data. Data upload followed quality and regulatory requirements already in place to ensure adequate uniformity and homogeneity levels. Data were formatted to meet the requirements of the FAIR Data Principles and thus made interoperable with other FAIR omics data and reference databases.

### Collected laboratory and instrumental data

A continuous quantitative respiratory score, the PaO_2_/FiO_2_ [Partial pressure of oxygen/Fraction of inspired oxygen ratio (P/F)] was assigned to each patient as an indicator of the respiratory involvement. Taking the normal value >300 as the threshold, we defined four grades of severity score for the PaO_2_/FiO_2_ ratio: P/F less than or equal to 100, between 101 and 200, between 201 and 300, and greater than 300. A P/F value is not available for the non-hospitalized subjects because the test is only performed in hospitalized patients when needed. Heart involvement was considered on the basis of one or more of the following abnormal data: a cardiac Troponin T (cTnT) value higher than the reference range (<15 ng/L) (indicative of ischemic disorder), an increase in the N-terminal (NT)-pro hormone BNP (NT-proBNP) value (reference value <88 pg/ml for males and <153 pg/ml for females) (indicative of heart failure), and the presence of arrhythmias (indicative of electric disorder). Hepatic involvement was defined on the basis of a clear liver enzymes elevation as alanine transaminase (ALT) and aspartate transaminase (AST) higher than the gender specific reference value (for ALT <41 UI/L in males and <31 UI/L in females; for AST <37 UI/L in males and <31 UI/L in females). Pancreatic involvement was considered on the basis of pancreatic enzymes as pancreatic amylase (PA) and lipase (PL) higher or lower than their specific reference range (13-53 UI/l for PA and 13-60UI/l per PL). Kidney involvement was defined in the presence of a creatinine value higher than the gender specific reference value (0,7-1,20 mg/dl in males and 0,5-1,10 mg/dl in females). Lymphoid system involvement was designated as Natural killer (NK) cells and/or peripheral CD4^+^ T cells below reference value (NK cells>90 cell/ul (mm ^^^3); CD4+T cells>400 cell/ul (mm^^^3)). For each patient a numerical grading for the olfactory and gustatory dysfunction was defined through a clinical questionnaire administered by ENT specialists. D-Dimer values of >10X, with or without low Fibrinogen level, were used to interpret the involvement of the blood clotting system. Interleukin 6 (IL6), lactate dehydrogenase (LDH), and c-reactive protein (CRP) values above the reference range (<0,5 mg/dl for CRP and 135-225 UI/l in males and 135-214 UI/l in females for LDH) were used to determine proinflammatory cytokines system involvement.

### Whole Exome sequencing

Whole Exome Sequencing with at least 97% coverage at 20x was performed using the Illumina NovaSeq6000 System (Illumina, San Diego, CA, USA). Library preparation was performed using the Illumina Exome Panel (Illumina) according to the manufacturer’s protocol. Library enrichment was tested by qPCR and the size distribution and concentration were determined using Agilent Bioanalyzer 2100 (Agilent Technologies, Santa Clara, CA, USA). The Novaseq6000 System (Illumina) was used for DNA sequencing through 150 bp paired-end reads.

### Genotyping

Genotyping data on 700,000 genetic markers were obtained on genomic DNA using the Illumina Global Screening Array (Illumina) according to the manufacturer’s protocol. Homo sapiens (human) Genome Reference Consortium Human Build 38 (GRCh38) was used. Quality checks (SNP calling quality, cluster separation, and Mendelian and replication error) were done using GenomeStudio analysis software (Illumina). The computer package Plink v1.90 [8] was used to process 700k SNP-genotyping data and to calculate SNP genotype statistics.

### Statistical analysis

Descriptive statistics were calculated to determine the distribution of clinical features by sex, age, and ethnicity. Chi-square tests were used to evaluate the statistical association between the clinical severity of the disease (from no hospitalization to intubation) and the categorical clinical variables: gender, ethnicity, blood group, respiratory severity, taste/smell involvement, heart involvement, liver involvement, pancreas involvement, kidney involvement, lymphoid involvement, cytokines trigger, D-dimer, and number of comorbidities. A linear regression model was used to test the statistical association between COVID-19 severity and age.

The variability within clinical features and their relative relationships have been summarised and described by principal component analysis (PCA). Only numerical variables with a missing rate lower than 50% were selected; these included hyposmia, neutrophils, CRP, fibrinogen, LDH, D-dimer, and number of comorbidities. Missing data were imputed using KNN (k-nearest neighbour) imputation [9], based on Gower distances [10]. After imputation, variables were centered and scaled prior to PCA. Descriptive statistics, chi-square tests, linear regression, and PCA were performed with the R environment for statistical computing [11].

A descriptive analysis of the phenotypes by using a hierarchically-clustered heatmap was performed. In particular, both patients and phenotypes are clusterized with the agglomerative hierarchical clustering methodology, where the chosen metric is the hamming distance and the linkage criterion is the “average” one (unweighted pair group method with arithmetic mean, UPGMA). The corresponding dendrograms of the clusterization are reported in the upper and in the left part of the heat plot. Then the information for the grading of severity of the patients is added a posteriori on the left strip. The resulting plot is obtained with the Python Seaborn package.

## RESULTS

The GEN-COVID Multicenter Study, through a cooperative and carefully curated moded of sample and data collection, has employed rigorous analyses to achieve phenotypic and genotypic data that can now be used to begin to identify host genetic dispositions to COVID-19. The careful methodological approach across a large geographical area to develop a biobank (the GCB), a registry (the GCPR), and finally the resulting genetic data collection (the GCGDC). Following the timelines and milestones of the GEN-COVID Multicenter Study (see **Figure 1**), the study has achieved a COVID-19 biobank, registry, and genetic data collection linked to one another, providing a high degree of confidence in sample and data integrity, and open to the world for COVID-19 research at what may still be considered an early point in this pandemic.

### The GEN-COVID Biobank (GCB)

The GEN-COVID Biobank (GCB), a collection of bio-specimens from patients affected by COVID-19 and the associated GEN-COVID Patient Registry (GCPR) were established and maintained at the University of Siena using the infrastructure of an already well-established biobank (est. 1998) (http://www.biobank.unisi.it/ScegliArchivio.asp).

The Biobank is closely linked to national and international biobanking efforts aimed at collecting high quality samples and patient data in a uniform manner and ensuring their FAIR (Findable, Accessible, Interoperable and Reusable) management. It is part of the BBMRI-IT [12], EuroBioBank (EBB; [13]), Telethon Network of Genetic Biobanks (TNGB; [14]), and RD-Connect [15]. The biobank and registry are ISO-certified (certificate 199556-2016-AQ-ITA-ACCREDIA) and accredited according to SIGU (the Italian Society of Human Genetics) requirements (Certificate 204107-2016-AQ-ITA-DNV).

Collected biological samples include peripheral blood, plasma, serum, primary leukocytes, and DNA samples. Samples were stored in a dedicated biobank section while associated clinical data were entered in the related registry. The biobank and registry were organized according to the highest scientific standards, preserving patients’ and citizens’ privacy, while providing services to the healthcare and scientific community to develop better treatments, test diagnostic tools, and advance COVID-19 and coronavirus research. Biobank personnel are responsible for sample pseudonymization, storage, and insertion in the online biobank catalogue.

### Geographical coverage

The GEN-COVID Multicenter Study reached a large number of subjects throughout Italy. Tuscany, which is the region in which the study is carried out, contributes presently 22.8% of enrolled patients. The Northern Italian regions, particularly Lombardy and Venetia, currently contribute 52.3% of enrolled patients (**Figure 2**). This distribution reflects closely the incidence of SARS-CoV-2 infection per 100,000 inhabitants for each Italian region, as updated to 4 July 2020 [3].

### The GEN-COVID Patient Registry (GCPR)

From April 7, 2020 to July 16, 2020, the GEN-COVID Patient Registry (GCPR) collected clinical data from a total of 1033 Italian SARS-Cov-2 PCR-positive individuals. For each individual, we collected clinical information using standardized clinical schedules (**Supplementary Table 1**). The study protocol also provides access to patients’ medical records and continual clinical data updating in order to secure continuity for patient follow-up.

The mean age of the entire cohort is presently 58.7 years (range 18-99). The cohort is presently predominantly male (57.1%) with a mean age of 59.5 years (range 18-99); the mean age of the females is 57.6 years (range 19-98) (**Table 1**). About 40.3% of the cohort has no chronic conditions. The overall case-fatality rate (CFR) is 3.6% (37 deaths among 1,033 cases with a mean age of 75.2 years [range 62-91]. Regarding the ethnicity, the cohort is composed of 998 White (96.61%), 21 Hispanic (2.03%), 4 Black (0.38%), and 10 Asian (0.96%) patients (**Table 1**).

**Table 1.** Characteristics of cohort.

|  |  |
| --- | --- |
| No. of subjects | 1,033 |
| Median age (range) <sup>a</sup> | 58,7 (18-99) |
| Gender No. (%) |  |
| *Male* | 590 (57,1%) |
| *Female* | 443 (42.9%) |
| Ethnicity No. (%) |  |
| *White* | 998 (96.61%) |
| *Hispanic* | 21 (2.03%) |
| *Black* | 4 (0.38%) |
| *Asian* | 10 (0.96%) |
| Clinical Category No. (%) |  |
| *Hospitalized intubated (Group 4)* | 98 (9.5%) |
| *Hospitalized CPAP/BiPAP (Group 3)* | 190 (18.4%) |
| *Hospitalize oxygen support (Group 2)* | 326 (31.55%) |
| *Hospitalized w/o oxygen support (Group 1)* | 153 (14.8%) |
| *Not hospitalized a/paucisymptomatic (Group 0)* | 266 (25.75%) |

Subjects have been divided into five qualitative severity clinical categories depending on the need for hospitalization, the respiratory impairment and, consequently, the type of ventilation required: i) hospitalized and intubated (9.5%); ii) hospitalized and CPAP-BiPAP and high-flows oxygen treated (18.4%); iii) hospitalized and treated with conventional oxygen support only (31.55%); iv) hospitalized without respiratory support (14.8%); v) not hospitalized pauci/asymptomatic individuals (25.75%) (Group 4 to 0 in **Table 1**).

Gender distribution was statistically significantly different among the 5 groups *(p-* value=7.81×10’^6^). In the group with high care intensity (Group 4), 72.4% of subjects were male, while in the group with the milder phenotype (Group 0) 59.8% of subjects were female (**Table 2**). Hyposmia and/or hypogeusia were present in 13.9% of cases in Group 4, 25.3% in Group 3, 31.6% in Group 2, 19.3% in Group 1, and in 57.1% of Group 0. A slight statistically significant difference among the 5 groups was found regarding the presence of comorbidities (p-value=0.012). No statistically significant difference was present for ethnicity and blood group distribution (**Table 2**).

**Table 2.** Cohort stratification by disease severity.

| **Subject characteristics** | **Group 4** | **Group 3** | **Group 2** | **Group 1** | **Group 0** | ***p-value*** |
| --- | --- | --- | --- | --- | --- | --- |
| **Median age (range) <sup>a</sup>** | 61.3 (29-79%) | 65.3 (21-91%) | 66.0 (21-99%) | 55.86 (25-93%) | 46.75 (19-72%) | 1.35x10<sup>-7</sup> |
| **Gender** |  |  |  |  |  |  |
| *Male (%)* | 72.4% | 71.5% | 59.8% | 53.6% | 40.2% | 7.81x10<sup>-6</sup> |
| *Female (%)* | 27.6% | 28.5% | 40.2% | 46.4% | 59.8% |  |
| **Ethnicity** |  |  |  |  |  |  |
| *White* | 97% | 97.3% | 96.7% | 96.6% | 99.6% | 0.731 |
| *Hispanic* | 2% | 1.1% | 1.5% | 2% | 0.4% |  |
| *Black* | 1% | 0 | 0.6% | 0.7% | 0 |  |
| *Asian* | 0 | 1.6% | 1.2% | 0,7% | 0 |  |
| **Blood Group** |  |  |  |  |  |  |
| *A* | 40.6% | 46.6% | 43.8% | 43.25% | 55% | 0.209 |
| *B* | 8.7% | 12.8% | 16.1% | 13.5% | 10% |  |
| *O* | 50.7% | 39.8% | 39.2% | 43.25% | 30% |  |
| *AB* | 0 | 0.8% | 0.9% | 0 | 5% |  |
| **Co-morbidities** |  |  |  |  |  |  |
| *None* | 21.7% | 18.8% | 26.3% | 32.4% | 72.5% | 0.012 |
| *One* | 34.8% | 30.1% | 26.7% | 35.1% | 17.5% |  |
| *More than one* | 42% | 45.1% | 45.6% | 28.4% | 10% |  |
| *Unknown* | 1.5% | 6% | 1.4% | 4.1% | 0 |  |

**Figure 3** shows the relationships between continually updated laboratory variables from PCA. The first two principal components explain 42.4% of the variability in the data (PC1: 23.8%; PC2: 18.6%). Neutrophils, LDH, and D-dimer appear to be positively correlated, while fibrinogen and CRP, and hyposmia and the number of comorbidities have been found, pairwise, to be negatively correlated. The largest contributors to PC1 were LDH (24.4%), neutrophils (24%), D-dimer (23.8%), and hyposmia (15.3%); the largest contributors to PC2 were hyposmia (25.8%), fibrinogen (22.1%), CRP (19.8%), and the number of comorbidities (15.4%) (**Figure 3**).

The continually updated laboratory values used in Figure 3 can be further mined through clinical reasoning and represented as a binary clinical classification for organ/system damage (**Table 3**).

**Table 3.** Binary clinical classification.

| **Organ /system** | **Value** | **Rule** | **Clinical Interpretation** |
| --- | --- | --- | --- |
| Lung | 1 , 0 | 1 if severity grading 4-2 and 0 if severity grading 1-0 | Lung disease |
| Heart | 1 , 0 | 1 if cTnT > reference value or NT-proBNP > gender specific reference value or Arrhythmia | Heart disease |
| Liver | 1 , 0 | 1 if ALT and AST > gender specific reference value | Liver disease |
| Pancreas | 1 , 0 | 1 if lipase and/or pancreatic amylase > or < specific reference value | Pancreas disease (either inflammation or depletion) |
| Kidney | 1 , 0 | 1 if creatinine > gender specific reference value | Kidney disease |
| Lymphoid system | 1 , 0 | 1 if NK cells < reference value or CD4 lymphocytes < reference value | Innate and adaptive immune deficit |
| Olfactory / gustatory system | 1 , 0 | 1 if Hypogeusia or Hyposmia | Olfactory and Gustatory deficit |
| Clotting system | 1 , 0 | 1 if D-dimer > 10X W/wo low Fibrinogen level (with high basal level) | Thromboembolism |
| Pro-inflammatory cytokines system | 1 , 0 | 1 if IL6 > reference value or LDH and CRP > reference value | Hyperinflammatory response |

cTnT, cardiac Troponin T; NT-proBNP, N-terminal (NT)-pro hormone BNP; ALT, Alanine transaminase; AST, Aspartate transaminase; CD4, CD4+ T cells; NK, Natural killer; IL6, Interleukin 6; LDH, Lactate dehydrogenase; CRP, c-reactive protein.

**Table 4** shows the prevalence of different organ/systems damage in the 5 different clinical categories based on respiratory failure (**Table 4**). Heart involvement was detected in 55% of subjects in Group 4, 39% of subjects in Group 3, 34.1% in Group 2, and 21.6% in Group 1. Liver involvement was present in 72.4% of cases in Group 4, 59.3% in Group 3, 46% in Group 2, and 33.7% in Group 1. Statistically significant difference among the 5 groups was found for all organs/systems, except for the lymphoid system.

**Table 4.** Cohort systemic description.

| **Organ/system Involvement** | **Group 0** | **Group 1** | **Group 2** | **Group 3** | **Group 4** | ***p-value*** |
| --- | --- | --- | --- | --- | --- | --- |
| Heart disease | 0* | 0,216 | 0,341 | 0,390 | 0,550 | 0.00016 |
| Liver disease | 0* | 0,337 | 0,460 | 0,593 | 0,724 | 2.96x10<sup>-33</sup> |
| Pancreas disease | 0* | 0,054 | 0,073 | 0,218 | 0,304 | 7.15x10<sup>-5</sup> |
| Kidney disease | 0* | 0,121 | 0,244 | 0,278 | 0,434 | 0.0117 |
| Innate and adaptive immune deficit | 0* | 0,202 | 0,138 | 0,270 | 0,507 | 0.229 |
| Olfactory / gustatory deficit | 0,4 | 0,162 | 0,225 | 0,157 | 0,086 | 0.0011 |
| Thromboembolism | 0* | 0,040 | 0,073 | 0,097 | 0,318 | 4.2x10<sup>-7</sup> |
| Hyperinflammatory response | NA | 0,081 | 0,152 | 0,278 | 0,492 | 2.02x10<sup>-5</sup> |

NA: not applicable, \*assigned on clinical ground;

Finally, **Figure 4** shows by dendrogram COVID-19 phenotype can be clustered using the above reported clinical data representations. Hierarchical Clustering analysis identified five main clinical categories and several subcategories: A) severe multisystemic, with either thromboembolic (A1) or pancreatic variant (A2); B) cytokine storm, either moderate (B1) or severe with liver involvement (B2); C) mild, either with (C1) or without hyposmia (C2); D) moderate, either without (D1) or with (D2) liver damage; E) heart type, either with (E1) or without (E2) liver damage (**Figure 4**).

### GEN-COVID Genetic Data Repository (GCGDR)

WES and Genotype (GWAS) data were generated within the GEN-COVID Genetic Data Repository (GCGDR). In order to be able to store and analyse the massive amount of genomic data (mainly WES with coverage > 97% at 20x, and prospetically including also WGS) generated with the analysis of the entire cohort of samples populating the biobank, we relied on the NIG. External users can upload and analyse data using the NIG pipeline by registering and creating a specific project. A section dedicated to COVID-19 samples has been created within the NIG database (http://nigdb.cineca.it/) that provides variant frequencies as a free tool for both clinicians and researchers.

The data from WES are available both in Variant Call Format (VCF) file or as binarized file, according to different classes of variants: i) rare variants (minor allele frequency (MAF)<1%); ii) low frequency variants (MAF<5%); iii) common polymorphisms (MAF>5%) in either homozygosity or supposed compound heterozygosity, with rare or low frequency variants. The distribution of these 3 classes of variants according to mutated genes in our cohort is shown in **Supplementary Figure 1**.

From WES 580,688 variants have been called: of these, 543,138 are SNP and 37,550 are MNP (multi-nucleotide polymorphisms). Exonic SNPs were distributed over the 22 autosomes of the human genome, plus the sex chromosomes. The average missing rate was 0.01, with per-sample maximum value of 0.017. 15,285 SNP loci had a missing rate greater than 5%. The average MAF was 0.032 (std. dev. 0.091), with a right-skewed distribution (median MAF = 0.0007). Only 1,041 SNPs were monomorphic (0.2%), but 437,246 (80.5%) had a frequency < 0.01. From the genotype perspective, the average observed heterozygosity was 0.047.

The data from high-density (700k) SNP genotyping are also generated on the same cohort and shared with international collaborations, including the COVID-19 Host Genetics Initiative (https://covid-19genehostinitiative.net/) and with GoFAIR VODAN [16]. From this analysis, SNP genotypes at 730,059 loci distributed over the entire human genome have been obtained. The average missing-rate was 0.015, with per-sample maximum value of 0.042. 11,163 SNP loci had a missing rate greater than 5%. The average MAF was 0.113 (std. dev. 0.145), with a right-skewed distribution (median MAF = 0.035). In total, 147,579 SNPs were monomorphic (20.2%). From the genotype perspective, the average observed heterozygosity was 0.155.

## DISCUSSION

The COVID-19 pandemic represents an enormous challenge for the world’s healthcare systems. The healthcare infrastructures and the working conditions are tremendously challenged in many hospitals and direct patient care has rightly been given the highest priority. The main public health focus is on monitoring infection evolution in terms of the number of new cases and the number of deaths as well as the number of patients experiencing serious pulmonary or systemic disease. To better characterize the current outbreak and facilitate prospective research to address the current and possible future epidemics/pandemics, we set up a COVID-19 biobank and patient registry where biological samples and associated clinical data from patients are collected in a standardized manner.

As expected, the majority of subjects in the group with high care intensity (Group 4) were males (72.4%) while in the group with mild phenotype the majority of subjects were females (59.2%). This is confirmatory of previously published data reporting a predominance of males among the most severely COVID-19 affected patients [17]. Among the 767 SARS-CoV-2-positive hospitalized patients in the current cohort, 63% are males and 12.8% required intubation. This is in line with the distribution of the Italian population of hospitalized COVID-19 patients [3] underlining the representativeness of our cohort.

Heart involvement was detected in the majority of severe cases (Group 4), confirming again a recent report [18]. Hospitalized SARS-CoV-2 positive patients (Group 2 to 4) have multiple-organ involvement: in particular, heart, liver, pancreas, and kidney. In line with our previous data and with literature findings, this confirms that COVID-19 is a systemic disease rather than simply a lung disorder [19;20].

### Clinical data representation and interpretation

Clinical data may be represented and consequently interpreted in different ways. The simplest way of representation is using the raw data of laboratory/instrument values. In this case, reasoning about which value has to be considered and/or at which time of clinical evolution the value needs to be measured is necessary in order to have consistency within the cohort. PCA analysis using the WORSEN score at the time of admission has shown the expected variability with hyposmia to be juxtaposed to the number of comorbidities and thus representing a marker of less severity. The fibrinogen value is juxtaposed to inflammatories markers, such as CRP (and D-Dimer and LDH) because it is consumed during the prothrombotic state. We can conclude that such raw laboratory values are fairly good for representing the clinical variability of the cohort in classical PCA analysis.

A more elaborate way of representing clinical data is to filter the raw laboratory/instrument values by clinical reasoning, which often requires a face-to-face meeting with organ reference specialists and direct access to the patients’ medical records. The proposed mediation of such a clinical methodology for COVID-19 is represented in **Table 3** and its distribution against lung dysfunction synthesised in **Table 4**.

Involvement of relevant organs or systems is represented in binary and is then used for representing COVID-19 as a systemic disorder (**Figure 4**). We propose this representation as one of the best, being closer to the real complexity of the disease. It should be considered for use in further data mining and correlation with genetic data. The emerging clinical categories from Hierarchical Cluster Analysis point to specific types and subtypes that are more likely to have common genetic factors.

As unmasked by our dendrogram (group A), there is indeed a growing body of evidence suggesting that, in addition to the common respiratory symptoms (fever, cough and dyspnea), COVID-19 severely-ill patients can often have symptoms of a multisystemic disorder [21]. Multiple organ failure due to diffuse microvascular damage is an important cause of death in COVID-19 severely affected patients [22]. In line with our definition of an A1 subgroup, a retrospective study on 21 deaths after SARS-Co-V2 infection recently reported that 71% of the patients who died had disseminated intravascular coagulation (DIC), while the incidence of DIC in surviving patients was 0.6% [23]. These data suggest that DIC is an important risk factor for increased in-hospital mortality and special attention should be paid to its early diagnosis and treatment.

While a debate still exists about the significance of pancreatic enzyme elevations during COVID-19 infection and the capability of SARS-CoV-2 virus to induce pancreatic injury due to cytotoxic effects [24, 25], it is worth noting that among patients with a multisystemic involvement we observe a subclass of individuals (group A2) with pancreatic damage, likely suggesting a secondary effect of SARS-CoV-2 infection on a subgroup of genetically predisposed individuals. Inflammatory cytokine “storm,” has been reported as playing a key role in the severe immune injury to the lungs caused by T-cell over-activation (group B) [26]. While some investigators have suggested a potential mechanism of myocardial injury due to COVID-19-induced cytokine storm that is mediated by a mixed T helper cell response in combination to hypoxia [27], our findings indicate rather a distinct class of patients (group E) presenting with heart involvement in the absence of an inflammatory cascade. This would tend to support the hypothesis that SARS-CoV-2 may directly damage myocardial tissue and induce a major cardiovascular event. Thus, as currently recommended, our research reinforces the need to monitor plasma cTnT and NT-proBNP levels in COVID-19 patients. In line with current evidence [28; 29], although liver injury seems to occur more frequently among critically ill patients with COVID-19 (group B), it can also be present in non-critically ill patients (groups D and E) and, as suggested, it could be mostly related to prolonged hospitalization and viral shedding duration. This allows defining, for each group, a clinical subclass according to this organ involvement.

A recent extensive review determined the prevalence of chemosensory deficits based on pooling together forty-two studies reporting on 23,353 patients [30]. Estimated random prevalence was 38.5% for olfactory dysfunction, 30.4% for taste dysfunction, and 50.2% for overall chemosensory dysfunction. No correlation with age was detected, but anosmia/hypogeusia decreased with disease severity and ethnicity turned out to play a significant role with Caucasians having a 3 to 6 times higher prevalence of chemosensory deficits than East Asians. In accordance with evidence found in the literature, hyposmia was mostly represented among patients in group C with mild clinical symptoms [31].

### Genetic data representation and interpretation

Similar to the clinical data, large aggregates of genetic data derived from WES may be represented, and consequently interpreted, in different ways. After variant calling, it is possible to use data as such, or variants can be prioritized and filtered according to standard bioinformatics procedures [32], such as damaging effect predictions, healthy population allele frequency, and gene constraints to variation.

Alternatively, it is also possible to represent data in a binary mode as follows: i) select missense, splicing, and loss of function variants below 1% (rare variants); ii) select missense, splicing, and loss of function variants between 1% and 5% (low frequency variants); iii) select missense, splicing, and loss of function variants above 5% (common polymorphisms) in either homozygosity or supposed compound heterozygosity with rare or low frequency variants. The majority of patients showed about 3% of mutated genes in the above class i), 5% in class ii) and 28% in class iii) variants (**Supplementary Figure 1A**). No patients showed variants in more than 8,000 genes (**Supplementary Figure 1B**).

Protein interaction network and pathway analysis have been widely used to uncover and describe genetic relationships in complex diseases, such as cancer [33;34]. For example, over-representation analysis of the biological processes and pathways significantly affected by mutations will be instrumental to empower the statistical detection of genetic signatures associated to specific COVID-19 phenotypes and to reduce the number of parameters to consider (e.g. dimensionality reduction) with the purpose of developing robust algorithms for the prediction of genetic susceptibility to COVID-19 infection and response. Variants, genes, or biological processes will be employed as features to train interpretable, supervised machine learning classifiers (e.g. gradient boosting decision trees [35;36]), which will ease the identification of the genetic factors associated with clinical phenotypes.

While data collection is being consolidated and brought to completion according to the study design, we have started to work on a relatively new methodology based on Topological Data Analysis to provide a detailed multidimensional and multiscale exploration of the whole exome data that can drive an AI selection of genes that provide higher predictive power in a machine learning model. The method will be presented, together with the results, in a forthcoming paper.

### Post-Mendelian model of complex diseases

Previous attempts to interpret the genetic bases of complex disorders have failed with very few exceptions, even in those disorders in which (like COVID-19) twin studies demonstrated a very high rate of heritability, such as in psychiatric disorders. The reason for this story containing such a lack of scientific success resides in several weak points in the overall genetic approach to complex diseases: i) the method used to represent the complexity of the phenotype; ii) the procedure employed to represent the huge amount of different genetic data; and iii) the absence of a robust mathematical model able to interpret genetic data in non-Mendelian (non-rare) disorders. This paper provides a contribution to the first 2 points, likely paving the way for a solution to the third.

Frequently the phenotype of common (complex) disorders is oversimplified, thus attenuating reliable correlation with genetic data. Limiting the representation to differences of single parameters, such as respiratory assistance (intubation, CPAP-BiPAP, oxygen supplementation, etc.), is a possible trap for studies on complex disorders, as may be the case with COVID-19. Similarly, genetic data are often too large to be mined and fragmented in different non-communicating methods, betting on either the power of common polymorphisms (GWAS) or the power of variant accumulation (burden gene test for WES). The binary representation we are proposing here, together with network propagation for feature reduction, and followed by machine learning approaches, may help in this task. A rare disorder called TAR (OMIM # 274000) is teaching us that combinatorial rules of rare variant(s) with more common polymorphism(s) is what we are looking for [37].

The GEN-COVID Multicenter Study with its Registry (GCPR), Biobank (GCB), and Genetic Data Repository (GCGDR) is structured to continually link with leading European and international research organizations, public and private, as well as with regulatory and public health authorities for developing COVID-19 and SARS-related medicines research and treatment protocols. The success of the developing research and understanding of COVID-19 and the underlying SARS-CoV-2 virus will rely in large part on human biological materials and patient-level data that is comprehensively collected and systematically organized with careful attention to sample and data integrity as well as the FAIR Data Principles. Improving diagnostics, developing existing or new therapeutics, improving treatment protocols, and even developing public health policies relies upon a foundation of evidence that requires the comprehensive, patient, and systematic collection and organizing of COVID-19 patient biological samples and data of high integrity, confidence, and interoperability. The GEN-COVID Multicenter Study’s GCPR, GCB, and GCGDR present a model that can be further explored as a systematic approach to sample and data collection while also being immediately deployable in our collective fight against COVID-19.

## Data Availability

The data and samples referenced here in the GEN-COVID Patient Registry and the GEN-COVID Biobank are available for consultation. You may contact the corresponding author, prof. Alessandra Renieri for further information.

http://nigdb.cineca.it/index.php

http://www.nig.cineca.it/

## ACKNOWLEDGEMENTS

This study is part of the GEN-COVID Multicenter Study, https://sites.google.com/dbm.unisi.it/gen-covid, the Italian multicenter study aimed to identify the COVID-19 host genetic bases. The *COVID-19 Biobank of Siena* is part of the *Genetic Biobank of Siena*, member of BBMRI-IT, of Telethon Network of Genetic Biobanks (project no. GTB18001), of EuroBioBank, and of D-Connect, provided us with specimens. We thank the CINECA consortium for providing computational resources and the Network for Italian Genomes NIG http://www.nig.cineca.it for its support. We thank private donors’ support to A.R. (Department of Medical Biotechnologies, University of Siena) for the COVID-19 host genetics research project (D.L n.18 of March 17, 2020). We also thank the COVID-19 Host Genetics Initiative (https://www.covid19hg.org/).

## ETHICS APPROVAL

The GEN-COVID study was approved by the University Hospital of Siena Ethical Review Board (Protocol n. 16929, dated March 16, 2020).

## AUTHOR CONTRIBUTIONS STATEMENT

EF, FM, AR and designed the study. CF and IM, were in charge of biological samples’ collection and biobanking. MB, FF were in charge of clinical data collection. MB, FF, AR, and FM performed analysis/interpretation of clinical data. AS and MB were in charge of DNA isolations from peripheral blood samples. FV, GD, AG, RT carried the sequencing experiments. EB, SF, FR, AS, FB, NP, MC, PP, SC, and MS performed bioinformatics and statistical analyses. SD, FC, and FF prepared Figures and Tables. SD, CF, AMP, FPC, AR, and EF wrote the manuscript. CF submitted this paper. All authors have reviewed and approved the manuscript.

## DATA AVAILABILITY AND DATA SHARING STATEMENT

The data and samples referenced here in the GEN-COVID Patient Registry and the GEN-COVID Biobank are available for consultation. You may contact the corresponding author, Prof. Alessandra Renieri for further information.

## ADDITIONAL INFORMATION

The authors declare no competing interests.

GEN-COVID Multicenter Study (https://sites.google.com/dbm.unisi.it/gen-covid)

Francesca Montagnani^3,12^, Laura Di Samo ^1^, Andrea Tommasi ^1,2^, Maria Palmieri ^1^, Susanna Croci^1^, Arianna Emiliozzi^3,12^, Massimiliano Fabbiani^12^, Barbara Rossetti^12^, Giacomo Zanelli^3,12^, Laura Bergantini^13^, Miriana D’Alessandro^13^, Paolo Cameli^13^, David Bennet^14^, Federico Anedda^14^, Simona Marcantonio^14^, Sabino Scolletta^14^, Federico Franchi^14^, Maria Antonietta Mazzei^15^, Susanna Guerrini^15^, Edoardo Conticini^16^, Luca Cantarini^16^, Bruno Frediani^16^, Danilo Tacconi^17^, Chiara Spertilli^17^, Marco Feri^18^, Alice Donati^18^, Raffaele Scala^19^, Luca Guidelli^19^, Genni Spargi^20^, Marta Corridi^20^, Cesira Nencioni^21^, Leonardo Croci^21^, Gian Piero Caldarelli^22^, Maurizio Spagnesi^23^, Paolo Piacentini^23^, Maria Bandini^23^, Elena Desanctis^23^, Silvia Cappelli^23^, Anna Canaccini^24^, Agnese Verzuri^24^, Valentina Anemoli^24^, Agostino Ognibene^25^, Massimo Vaghi^26^, Antonella D’Arminio Monforte^27^, Esther Merlini^27^, Mario U. Mondelli^28,29^, Stefania Mantovani^28^, Serena Ludovisi^28,29^, Massimo Girardis^30^, Sophie Venturelli^30^, Marco Sita^30^, Andrea Cossarizza^31^, Andrea Antinori^32^, Alessandra Vergori^32^, Stefano Rusconi^33,34^, Matteo Siano^34^, Arianna Gabrieli^34^, Agostino Riva^33,34^, Daniela Francisci^35,36^, Elisabetta Schiaroli^35^, Pier Giorgio Scotton^37^, Francesca Andretta^37^, Sandro Panese^38^, Renzo Scaggiante^39^, Francesca Gatti^39^, Saverio Giuseppe Parisi^40^, Francesco Castelli^41^, Maria Eugenia Quiros-Roldan^41^, Paola Magro^41^, Isabella Zanella^42^, Matteo Della Monica^43^, Carmelo Piscopo^43^, Mario Capasso^44,45,46^, Roberta Russo^44,45^, Immacolata Andolfo^44,45^, Achille Iolascon^44,45^, Giuseppe Fiorentino^47^, Massimo Carella^48^, Marco Castori^48^, Giuseppe Merla^48^, Filippo Aucella^49^, Pamela Raggi^50^, Carmen Marciano^50^, Rita Perna^50^, Matteo Bassetti^51,52^, Antonio Di Biagio^52^, Maurizio Sanguinetti^53,54^, Luca Masucci^53,54^, Chiara Gabbi^55^, Serafina Valente^56^, Ilaria Meloni, Maria Antonietta Mencarelli, Caterina Lo Rizzo, Elena Bargagli, Marco Mandalà, Alessia Giorli^57^, Lorenzo Salerni^57^, Patrizia Zucchi^58^, Pierpaolo Parravicini^58^, Elisabetta Menatti^59^, Stefano Baratti^60^, Tullio Trotta^61^, Ferdinando Giannattasio^61^, Gabriella Coiro^61^, Fabio Lena^62^, Domenico A. Coviello^63^, Cristina Mussini^64^, Giancarlo Bosio^65^, Sandro Mancarella^66^, Luisa Tavecchia^66^.

12)Dept of Specialized and Internal Medicine, Tropical and Infectious Diseases Unit

13)Unit of Respiratory Diseases and Lung Transplantation, Department of Internal and Specialist Medicine, University of Siena

14)Dept of Emergency and Urgency, Medicine, Surgery and Neurosciences, Unit of Intensive Care Medicine, Siena University Hospital, Italy

15)Department of Medical, Surgical and Neuro Sciences and Radiological Sciences, Unit of Diagnostic Imaging, University

16)Rheumatology Unit, Department of Medicine, Surgery and Neurosciences, University of Siena, Policlinico Le Scotte, Italy

17)Department of Specialized and Internal Medicine, Infectious Diseases Unit, San Donato Hospital Arezzo, Italy

18)Dept of Emergency, Anesthesia Unit, San Donato Hospital, Arezzo, Italy

19)Department of Specialized and Internal Medicine, Pneumology Unit and UTIP, San Donato Hospital, Arezzo, Italy

20)Department of Emergency, Anesthesia Unit, Misericordia Hospital, Grosseto, Italy

21)Department of Specialized and Internal Medicine, Infectious Diseases Unit, Misericordia Hospital, Grosseto, Italy

22)Clinical Chemical Analysis Laboratory, Misericordia Hospital, Grosseto, Italy

23)Department of Preventive Medicine, Azienda USL Toscana Sud Est, Italy

24)Territorial Scientific Technician Department, Azienda USL Toscana Sud Est, Italy

25)Clinical Chemical Analysis Laboratory, San Donato Hospital, Arezzo, Italy

26)Chirurgia Vascolare, Ospedale Maggiore di Crema, Italy

27)Department of Health Sciences, Clinic of Infectious Diseases, ASST Santi Paolo e Carlo, University of Milan, Italy

28)Division of Infectious Diseases and Immunology, Fondazione IRCCS Policlinico San Matteo, Pavia, Italy

29)Department of Internal Medicine and Therapeutics, University of Pavia, Italy

30)Department of Anesthesia and Intensive Care, University of Modena and Reggio Emilia, Modena, Italy

31)Department of Medical and Surgical Sciences for Children and Adults, University of Modena and Reggio Emilia, Modena, Italy

32)HIV/AIDS Department, National Institute for Infectious Diseases, IRCCS, Lazzaro Spallanzani, Rome, Italy

33)III Infectious Diseases Unit, ASST-FBF-Sacco, Milan, Italy

34)Department of Biomedical and Clinical Sciences Luigi Sacco, University of Milan, Milan, Italy

35)Infectious Diseases Clinic, Department of Medicine 2, Azienda Ospedaliera di Perugia and University of Perugia, Santa Maria Hospital, Perugia, Italy

36)Infectious Diseases Clinic, “Santa Maria” Hospital, University of Perugia, Perugia, Italy

37)Department of Infectious Diseases, Treviso Hospital, Local Health Unit 2 Marca Trevigiana, Treviso, Italy

38)Clinical Infectious Diseases, Mestre Hospital, Venezia, Italy.

39)Infectious Diseases Clinic, ULSS1, Belluno, Italy

40)Department of Molecular Medicine, University of Padova, Italy

41)Department of Infectious and Tropical Diseases, University of Brescia and ASST Spedali Civili Hospital, Brescia, Italy

42)Department of Molecular and Translational Medicine, University of Brescia, Italy; Clinical Chemistry Laboratory, Cytogenetics and Molecular Genetics Section, Diagnostic Department, ASST Spedali Civili di Brescia, Italy

43)Medical Genetics and Laboratory of Medical Genetics Unit, A.O.R.N. “Antonio Cardarelli”, Naples, Italy

44)Department of Molecular Medicine and Medical Biotechnology, University of Naples Federico II, Naples, Italy

45)CEINGE Biotecnologie Avanzate, Naples, Italy

46)IRCCS SDN, Naples, Italy

47)Unit of Respiratory Physiopathology, AORN dei Colli, Monaldi Hospital, Naples, Italy

48)Division of Medical Genetics, Fondazione IRCCS Casa Sollievo della Sofferenza Hospital, San Giovanni Rotondo, Italy

49)Department of Medical Sciences, Fondazione IRCCS Casa Sollievo della Sofferenza Hospital, San Giovanni Rotondo, Italy

50)Clinical Trial Office, Fondazione IRCCS Casa Sollievo della Sofferenza Hospital, San Giovanni Rotondo, Italy

51)Department of Health Sciences, University of Genova, Genova, Italy

52)Infectious Diseases Clinic, Policlinico San Martino Hospital, IRCCS for Cancer Research Genova, Italy

53)Microbiology, Fondazione Policlinico Universitario Agostino Gemelli IRCCS, Catholic University of Medicine, Rome, Italy

54)Department of Laboratory Sciences and Infectious Diseases, Fondazione Policlinico Universitario A. Gemelli IRCCS, Rome, Italy

55)Independent Scientist, Milan, Italy

56)Department of Cardiovascular Diseases, University of Siena, Siena, Italy

57)Otolaryngology Unit, University of Siena, Italy

58)Department of Internal Medicine, ASST Valtellina e Alto Lario, Sondrio, Italy

59)Study Coordinator Oncologia Medica e Ufficio Flussi Sondrio, Italy

60)Department of Infectious and Tropical Diseases, University of Padova, Padova, Italy

61)First Aid Department, Luigi Curto Hospital, Polla, Salerno, Italy

62)Local Health Unit-Pharmaceutical Department of Grosseto, Toscana Sud Est Local Health Unit, Grosseto, Italy

63)U.O.C. Laboratorio di Genetica Umana, IRCCS Istituto G. Gaslini, Genova, Italy.

64)Infectious Diseases Clinics, University of Modena and Reggio Emilia, Modena, Italy.

65)Department of Respiratory Diseases, Azienda Ospedaliera di Cremona, Cremona, Italy

66)U.O.C. Medicina, ASST Nord Milano, Ospedale Bassini, Cinisello Balsamo (MI), Italy

**Figure 1.**
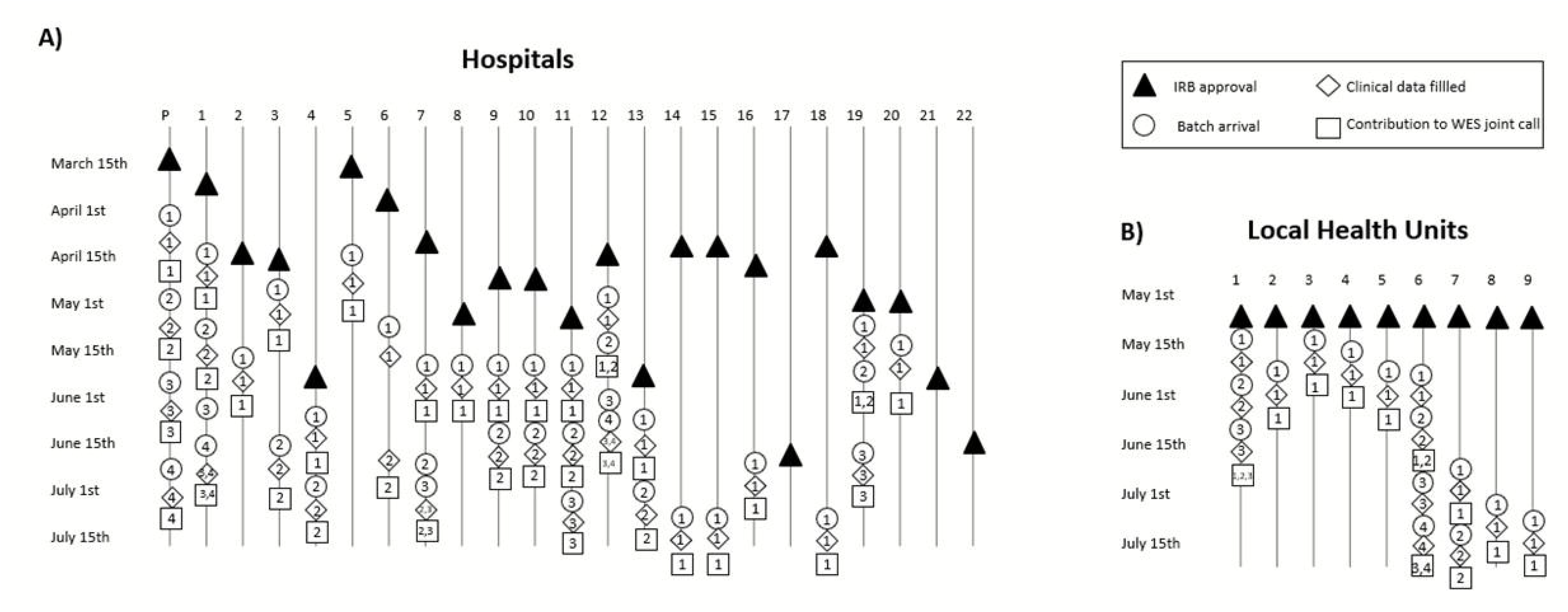
Timeline of GEN-COVID Multicenter study. **Panel A**. Main milestones of the study with the timeline for the 22 Italian hospitals (P: Promoter, Policlinico Santa Maria Alle Scotte, Azienda Ospedaliera Universitaria Senese, Siena; 1: San Matteo Hospital Fondazione IRCCS, Pavia; 2:ASST Santi Paolo e Carlo, University of Milan, Italy; 3: Ospedale Maggiore di Crema, Italy; 4: ASST Valtellina e Alto Lario, Sondrio; 5: University Hospital of Modena and Reggio Emilia, Modena; 6: IRCCS, Lazzaro Spallanzani, Rome; 7: ASST-FBF-Sacco, Milan; 8: Santa Maria Hospital, Azienda Ospedaliera di Perugia, Perugia; 9: Treviso Hospital, Local Health Unit (ULSS) 2 Marca Trevigiana, Treviso; 10: Ospedale dell’Angelo, ULSS 3 Serenissima, Mestre; 11: Belluno Hospital, ULSS 1 Dolomiti, Belluno; 12: ASST Spedali Civili Hospital, Brescia; 13: Policlinico San Martino Hospital, IRCCS, Genova; 14: AORN dei Colli, Monaldi Hospital, Naples; 15: A.O.R.N. “Antonio Cardarelli”, Naples; 16: Fondazione IRCCS Casa Sollievo della Sofferenza Hospital, San Giovanni Rotondo; 17: IRCCS Istituto G. Gaslini, Genoa; 18: CEINGE Biotecnologie Avanzate, Naples; 19: San Donato Hospital, Arezzo; 20: Misericordia Hospital, Grosseto; 21: Fondazione Policlinico Universitario Agostino Gemelli IRCCS; 22: Luigi Curto Hospital, Polla (SA). **Panel B**. Main milestones of the study with the timeline for local health units (Continuity Assistance Special Units, USCA) and departments of preventive medicine (1. USCA, Chianciano; 2: USCA Sansepolcro; 3: USCA Siena; 4: USCA Orbetello; 5: USCA Arezzo; 6: Department of preventive medicine Senese, Siena; 7: Department of preventive medicine Aretino-Casentino-Valtiberina, Arezzo; 8: Department of preventive medicine Alta Val d’Elsa, Poggibonsi; 9: Department of preventive medicine Amiata Senese e Val d’Orcia - Valdichiana Senese, Montepulciano). Other 11 USCA and 4 departments of preventive medicine have obtained IRB approval and they are going to start sample collection.

**Figure 2.**
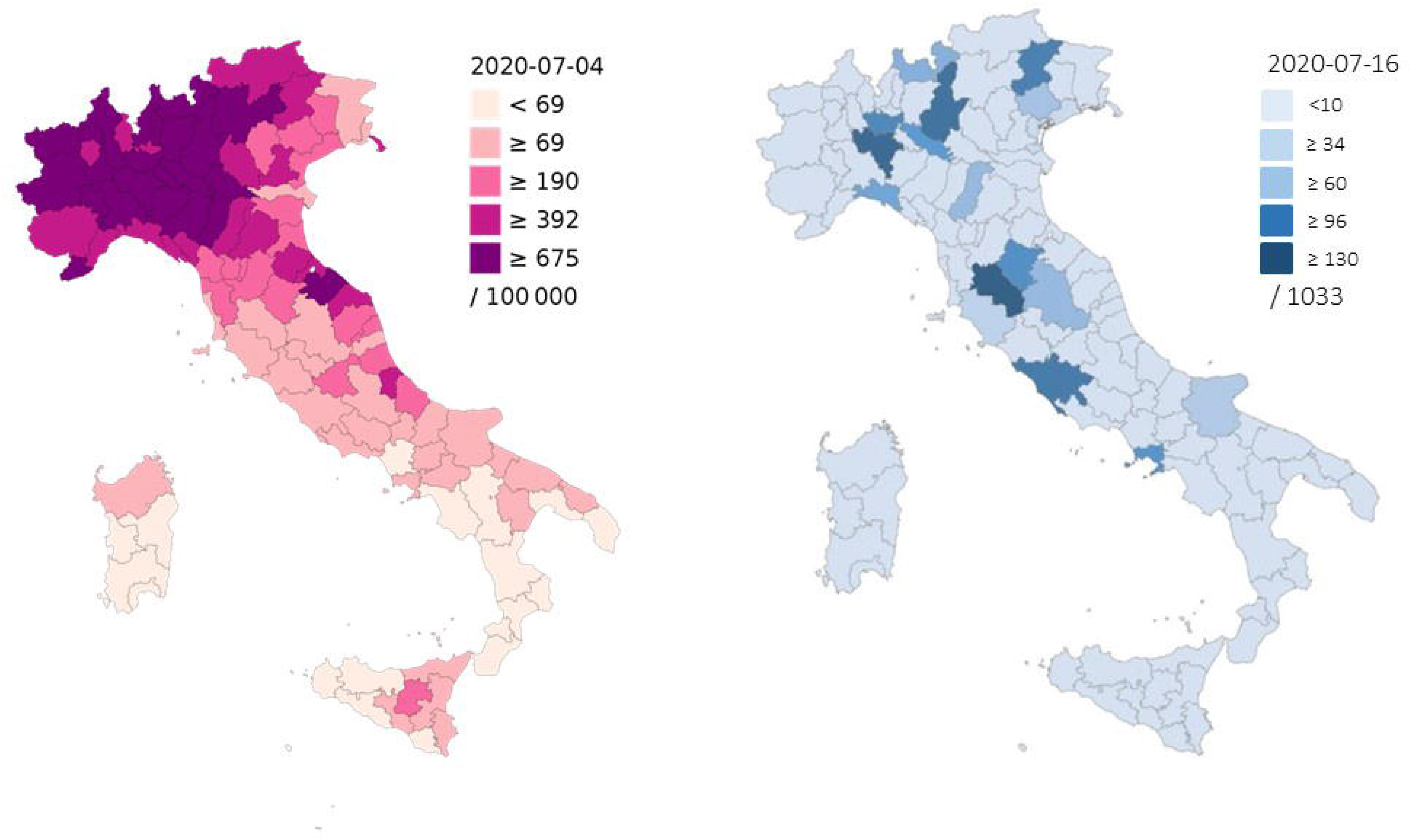
Geographical coverage. Comparison of GEN-COVID geographical coverage (right) and the incidence of SARS-CoV-2 infection per 100,000 inhabitants by Italian provinces (left).

**Figure 3.**
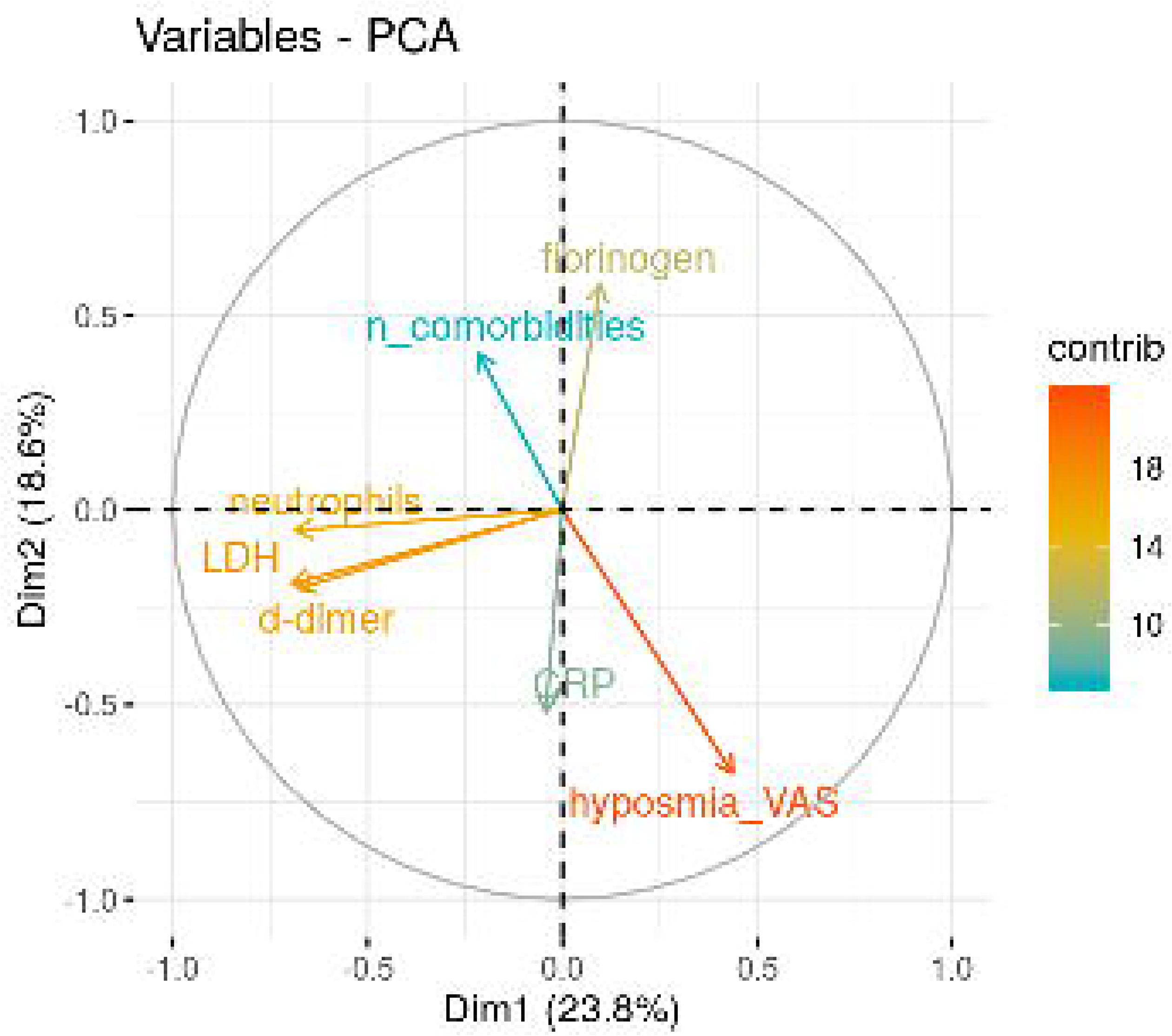
PCA variables plot. Eigenvector-based coordinates of the original variables in the two-dimensional space defined by the first two principal components. The relative position of the clinical variables reflect their relationship (positive correlated variables point to the same side of the plot; negative correlated variables point to opposite sides of the plot), while the length of the arrow is proportional to their contribution to the principal components.

**Figure 4.**
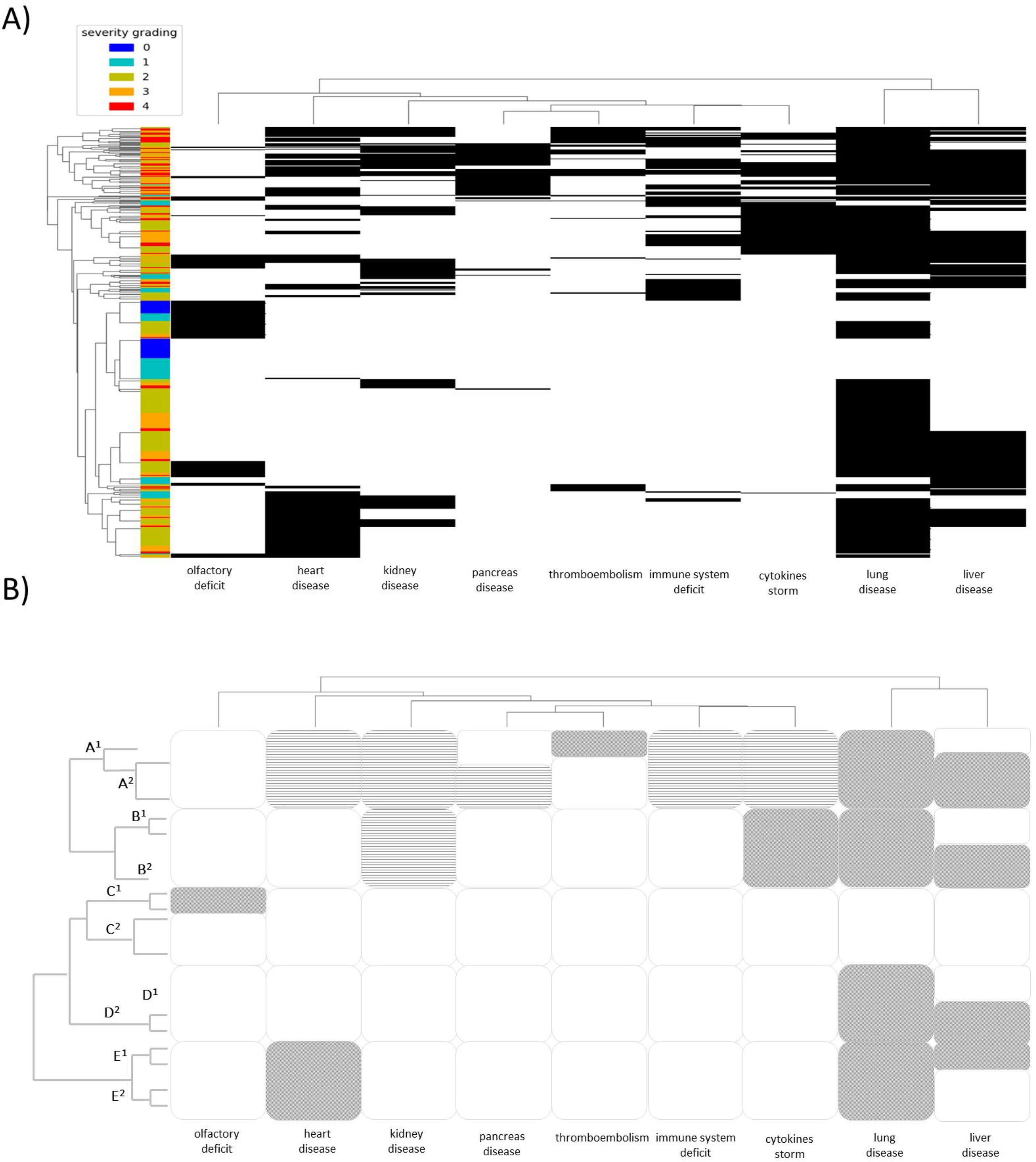
Phenotypic Clustering of COVID-19 patients. **Panel A**. Dendrogram of COVID-19 patients’ clinical phenotypes by hierarchical clustering of organ/system involvement. **Panel B**. Drawing of the above reported graph helping interpretation and simplification in the main branch of the tree. A^1^ severe multisystemic with either thromboembolic; A^2^ severe multisystemic with pancreatic variant; B^1^ cytokine storm with moderate liver involvement; B^2^ cytokine storm with severe liver involvement; C^1^ mild either with hyposmia; C^2^ mild without hyposmia; D^1^ moderate without liver damage; D^2^ moderate with liver damage; E^1^ heart with liver damage; E^2^ heart without liver damage.

**Supplementary Figure 1. Binary representation of WES data.**

Variants are divided in three classes: rare (minor allele frequency, MAF < 1%), low frequency (MAF < 5%) and common (MAF >5%). Panel A. Percentage of mutated genes in the three classes of variants. Panel B. Percentage of patients with mutated genes.

## REFERENCES

1. Wilkinson, M., Dumontier, M., Aalbersberg, I. et al. The FAIR Guiding Principles for scientific data management and stewardship. Sci Data 3, 160018 (2016). https://doi.org/10.1038/sdata.2016.18

2. Zhu N, Zhang D, Wang W, Li X, Yang B, Song J, et al. A Novel Coronavirus from Patients with Pneumonia in China, 2019. N Engl J Med. 2020;382:727–33.

3. Italian Civil Protection Department - COVID-19 case update (http://opendatadpc.maps.arcgis.com/apps/opsdashboard/index.htmltf/b0c68bce2cce478eaac82fe38d4138b1)

4. Wu Z, McGoogan JM. Characteristics of and important lessons from the coronavirus disease 2019 (COVID-19) outbreak in China: summary of a report of 72 314 cases from the Chinese Center for Disease Control and Prevention. JAMA. 2020 February 24. doi: 10.1001/jama.2020.2648.

5. Benetti E, Tita R, Spiga O, et al. ACE2 gene variants may underlie interindividual variability and susceptibility to COVID-19 in the Italian population [published online ahead of print, 2020 Jul 17]. Eur J Hum Genet. 2020;1-13.

6. Regulation (EU) 2016/679 of the European Parliament and of the Council of 27 April 2016 on the protection of natural persons with regard to the processing of personal data and on the free movement of such data, and repealing Directive 95/46/EC (General Data Protection Regulation).

7. Benetti E, Giliberti A, Emiliozzi A, Valentino F, Bergantini L, Fallerini C, et al. Clinical and molecular characterization of COVID-19 hospitalized patients. 2020. http://medrxiv.org/content/early/2020/05/25/2020.05.22.20108845

8. Chang, C.C., Chow, C.C., Tellier, L.C., Vattikuti, S., Purcell, S.M. and Lee, J.J., Second-generation PLINK: rising to the challenge of larger and richer datasets. Gigascience, 2015, 4(1), pp.s13742-015.

9. Troyanskaya, O., Cantor, M., Sherlock, G., Brown, P., Hastie, T., Tibshirani, R., et al. Missing value estimation methods for DNA microarrays. Bioinformatics, 2001, 17(6):520-525.

10. Gower JC. A general coefficient of similarity and some of its properties. Biometrics. 1971;27(4):857.

11. R Core Team. R: A language and environment for statistical computing. 2018, R Foundation for Statistical Computing, Vienna, Austria.

12. ‘BBMRI.it’. https://www.bbmri.it/ (accessed Mar. 30, 2020).

13. ‘EuroBioBank – EuroBioBank website’. http://www.eurobiobank.org/ (accessed Mar. 30, 2020).

14. ‘Telethon Network of Genetic Biobanks’. http://biobanknetwork.telethon.it/ (accessed Mar. 30, 2020).

15. ‘RD-Connect – RD-Connect website’. https://rd-connect.eu/ (accessed Mar. 30, 2020).

16. COVID-19 Host Genetics Initiative. The COVID-19 Host Genetics Initiative, a global initiative to elucidate the role of host genetic factors in susceptibility and severity of the SARS-CoV-2 virus pandemic. Eur J Hum Genet. 2020;28(6):715–718. doi:10.1038/s41431-020-0636-6

17. Cai, H. Sex difference and smoking predisposition in patients with COVID-19. The Lancet Respiratory Medicine. 2020, 8.4: e20.

18. Knight DS, Kotecha T, Razvi Y, Chacko L, Brown JT, Jeetley PS, et al. COVID-19: Myocardial injury in survivors. Circulation. 2020, Jul 14. doi: 10.1161/CIRCULATIONAHA.120.049252.

19. Waisse S, Oberbaum M, Frass M. The Hydra-Headed Coronaviruses: Implications of COVID-19 for Homeopathy. Homeopathy. 2020 Jul 22. doi:0.1055/s-0040-1714053.

20. Massabeti R, Cipriani MS, Valenti I. Covid-19: A systemic disease treated with a wide-ranging approach: A case report. J Popul Ther Clin Pharmacol, 2020 Jul 03, 27(SPt1):e26-e30.

21. Zheng KI, Feng G, Liu WY, Targher G, Byrne CD, Zheng MH. Extrapulmonary complications of COVID-19: A multisystem disease? [published online ahead of print, 2020 Jul 10]. J Med Virol. 2020;10.1002/jmv.26294. doi:10.1002/jmv.26294

22. Chen G, Wu D, Guo W, et al. Clinical and immunological features of severe and moderate coronavirus disease 2019. J Clin Invest. 2020, 130(5):2620-2629

23. Tang N, Li D, Wang X, Sun Z. Abnormal coagulation parameters are associated with poor prognosis in patients with novel coronavirus pneumonia. J Thromb Haemost. 2020, 18(4):844-847

24. Wang F, Wang H, Fan J, et al. Pancreatic injury patterns in patients with COVID-19 pneumonia. Gastroenterology 2020; Ashok A, Faghih M, Singh VK. Mild Pancreatic Enzyme Elevations in COVID-19 Pneumonia: Synonymous with Injury or Noise? [published online ahead of print, 2020 Jun 13]. Gastroenterology. 2020, S0016-5085(20)34778-8. doi:10.1053/j.gastro.2020.05.086

25. Szatmary P, Arora A, Raraty MGT, Dunne DFJ, Baron RD, Halloran CM. Emerging phenotype of SARS-CoV2 associated pancreatitis. Gastroenterology. 2020, Jun 1;S0016-5085(20)34741-7

26. Xu Z, Shi L, Wang Y, et al. Pathological findings of COVID-19 associated with acute respiratory distress syndrome. Lancet Respir Med. 2020, 8(4):420-422

27. Zheng YY, Ma YT, Zhang JY, Xie X. COVID-19 and the cardiovascular system. Nat Rev Cardiol. 2020, 17(5):259-260

28. Jiang S, Wang R, Li L, Hong D, Ru R, Rao Y, et al. Liver Injury in Critically Ill and Non-critically Ill COVID-19 Patients: A Multicenter, Retrospective, Observational Study. Front Med (Lausanne). 2020 Jun 23;7:347.doi: 10.3389/fmed.2020.00347. eCollection 2020.

29. Feng G, Zheng KI, Yan QQ, Rios RS, Targher G, Byrne CD, et al. COVID-19 and Liver Dysfunction: Current Insights and Emergent Therapeutic Strategies. J Clin Transl Hepatol. 2020 Mar 28;8(1):18–24.doi: 10.14218/JCTH.2020.00018. Epub 2020 Mar 30.

30. Von Bartheld CS, Hagen MM, Butowt R. Prevalence of Chemosensory Dysfunction in COVID-19 Patients: A Systematic Review and Meta-analysis Reveals Significant Ethnic Differences. Preprint. medRxiv. 2020, 2020.06.15.20132134. Published 2020 Jun 17. doi:10.1101/2020.06.15.20132134

31. Kim GU, Kim MJ, Ra SH, et al. Clinical characteristics of asymptomatic and symptomatic patients with mild COVID-19. Clin Microbiol Infect. 2020;26(7):948.e1-948.e3. doi:10.1016/j.cmi.2020.04.040

32. Eilbeck K, Quinlan A, Yandell M. Settling the score: variant prioritization and Mendelian disease. Nat Rev Genet. 2017 Oct;18(10):599-612.doi: 10.1038/nrg.2017.52. Epub 2017 Aug 14

33. Creixell P, Reimand J, Haider S, Wu G, Shibata T, Vazquez M, et al. Pathway and network analysis of cancer genomes. Nat Methods. 2015 Jul;12(7):615-621.doi: 10.1038/nmeth.3440.

34. Cowen L, Ideker T, Raphael BJ, Sharan R. Network propagation: a universal amplifier of genetic associations. Nat Rev Genet. 2017 Sep;18(9):551-562.doi: 10.1038/nrg.2017.38. Epub 2017 Jun 12.

35. Yan L, Zhang H-T, Goncalves J, Xiao Y, Wang M, Guo Y, et al. An interpretable mortality prediction model for COVID-19 patients. Nat Mach Intell 2, 2020; 283–288. https://doi.org/10.1038/s42256-020-0180-7

36. Shuai S, PCAWG Drivers and Functional Interpretation Working Group; Steven Gallinger, Lincoln Stein, PCAWG Consortium. Combined burden and functional impact tests for cancer driver discovery using DriverPower. Nat Commun. 2020 Feb 5;11(1):734.doi: 10.1038/s41467-019-13929-1.

37. Albers CA, Paul DS, Schulze H, et al. Compound inheritance of a low-frequency regulatory SNP and a rare null mutation in exon-junction complex subunit RBM8A causes TAR syndrome. Nat Genet. 2012;44(4):435-S2. Published 2012 Feb 26. doi: 10.1038/ng.1083

